# Combination therapy of infliximab and thiopurines, but not monotherapy with infliximab or vedolizumab, is associated with attenuated IgA and neutralisation responses to SARS-CoV-2 in inflammatory bowel disease

**DOI:** 10.1101/2021.10.13.21264916

**Authors:** Judith Wellens, Matthew Edmans, Uri Obolski, Colleen GC McGregor, Peter Simmonds, Marc Turner, Lisa Jarvis, Donal Skelley, Susanna Dunachie, Eleanor Barnes, David W Eyre, Jean-Frederic Colombel, Serre-Yu Wong, Paul Klenerman, James O Lindsey, Jack Satsangi, Craig P Thompson

## Abstract

There is substantial interest regarding the perceived risk that immunomodulator and biologic therapy could have on COVID-19 disease severity among patients with inflammatory bowel disease (IBD) and clinicians. In this study, we show that infliximab/thiopurine combination therapy is associated with significantly lower IgA, a range of lower IgG responses as well as impaired neutralising antibody responses, compared to responses observed in healthy individuals. We also demonstrate that whilst IgG responses were significantly reduced in individuals with IBD treated with infliximab or vedolizumab monotherapy compared to healthy controls, there was no significant reduction in IgA and neutralising antibody responses. As neutralising antibody responses correlate with protection, this observation may provide the mechanistic explanation for the observation reported by the SECURE-IBD study that individuals on infliximab/thiopurine combination therapy were at greater risk of severe COVID-19 outcomes than patients on monotherapy.

## Text

The effect of immunomodulator and biological therapy for inflammatory bowel disease (IBD) on the immune response to SARS-CoV-2 is of substantial interest to patients and clinicians worldwide. The CLARITY IBD study recently reported attenuated serological responses in IBD patients treated with infliximab in comparison to vedolizumab^1^, with the effect greatest in those on infliximab/thiopurine combination therapy. Independently, the global SECURE-IBD registry highlighted that infliximab/thiopurine combination therapy, but not infliximab or vedolizumab monotherapies, was associated with more severe clinical outcomes upon SARS-CoV-2 infection^2,3^.

However, these studies have not addressed treatment effects on neutralising antibody responses, the key correlate of protection to SARS-CoV-2; nor have they analysed the range of serological signatures that may influence clinical outcomes^4,5^.

To answer these questions, we performed an extended analysis of serological responses to SARS-CoV-2 infection in seropositive IBD patients treated with either infliximab or vedolizumab monotherapy, or infliximab/thiopurine combination therapy (Figures 1&2). Blood samples were collected from consenting patients attending infusion centres in Oxford and London between May and December 2020. Sera were initially screened by Abbott assay for SARS-CoV-2 antibody responses^6^. Serological reactivity profiles in positive samples were compared with those from healthy adult controls seropositive in the same assay^7^ (Supplementary information table 1).

**Figure 1.**
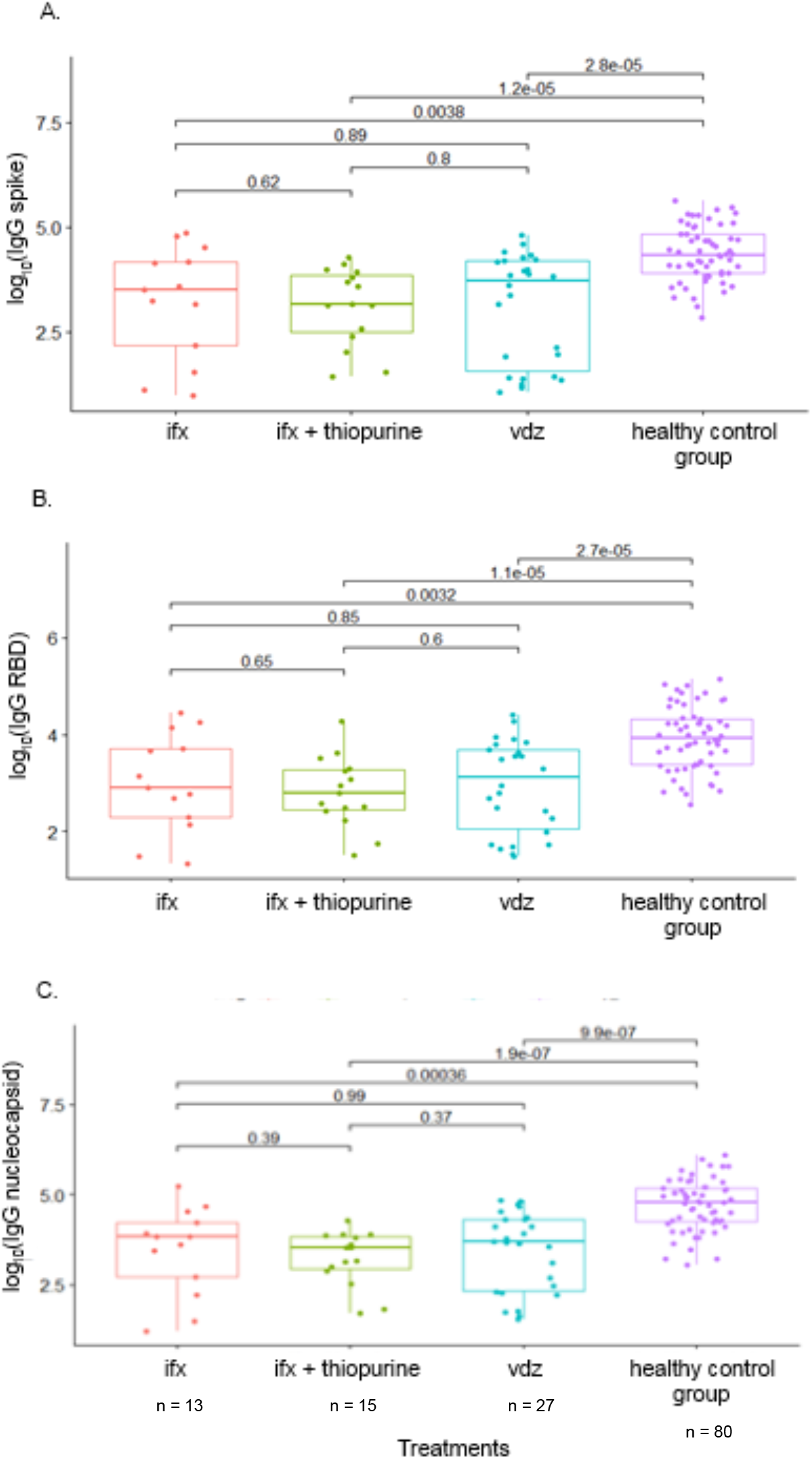
IgG responses to whole spike, receptor binding domain and nucleocapsid following SARS-CoV2 detection in IBD patients and healthy controls. A. IgG SARS-CoV-2 spike responses measured by high throughput V-PLEX MSD ELISA^8^. B. IgG SARS-CoV-2 receptor binding domain (RBD) of the spike responses measured by VPLEX MSD. C. IgG SARS-CoV-2 nucleocapsid responses measured by VPLEX MSD. Ifx = infliximab monotherapy, ifx+ thiopurines = infliximab/thiopurine combination therapy vdz = vedolimamab monotherapy. P-values are derived from a Wilcoxon (rank-sum) test for unpaired populations, not adjusted for multiple comparisons.

**Figure 2.**
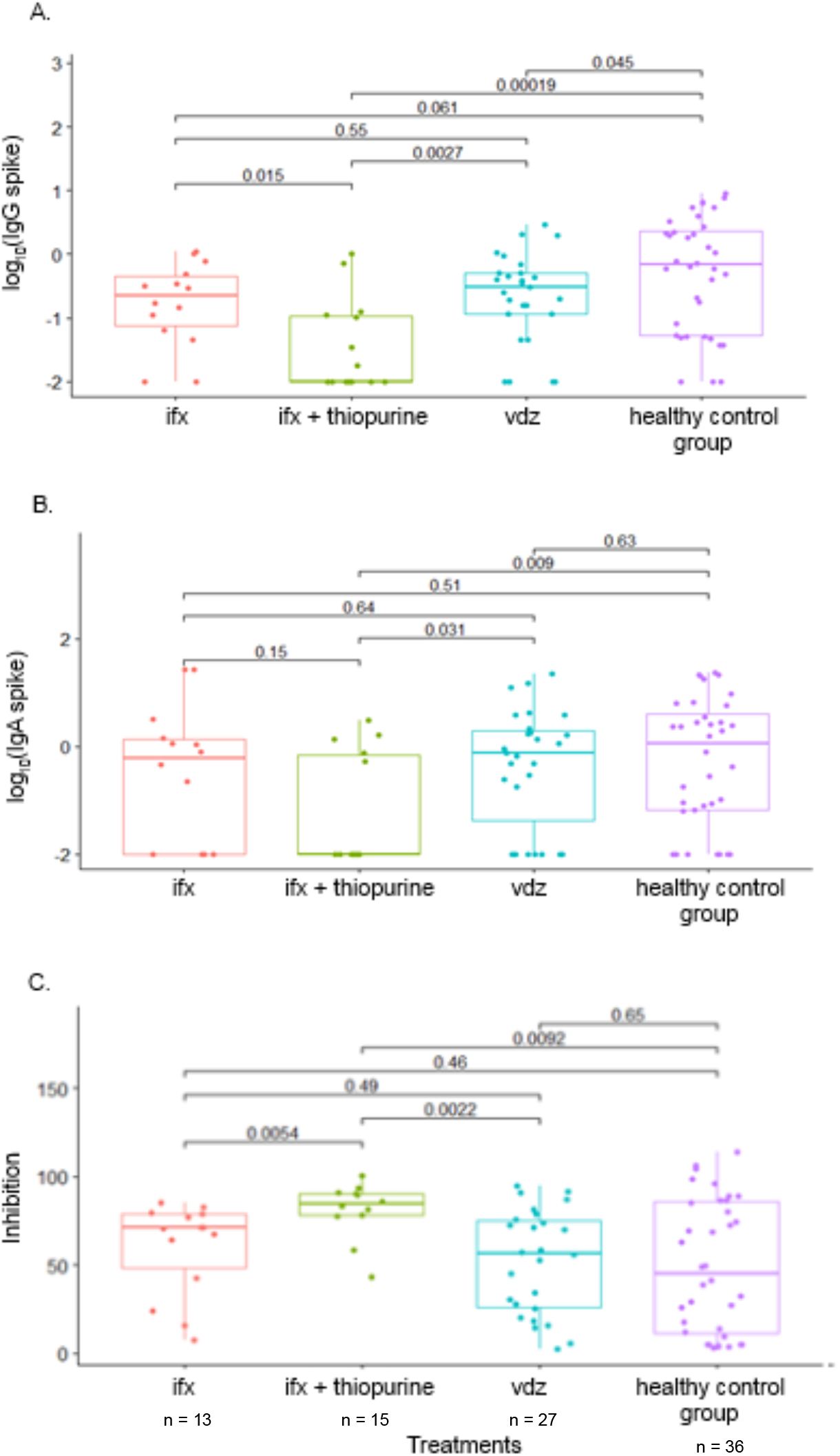
Neutralisation and IgA/IgG response following SARS-CoV2 detection in IBD patients and healthy controls. A. IgG SARS-CoV-2 spike responses measured by indirect ELISA. B IgA SARS-CoV-2 spike responses measured by indirect ELISA. C. Neutralising antibody responses measured by ACE2-RBD inhibition ELISA. Please note, higher responses indicate greater neutralisation. Ifx = infliximab monotherapy, ifx+ thiopurines = infliximab/thiopurine combination therapy vdz = vedolimamab monotherapy. P-values are derived from a Wilcoxon (rank-sum) test for unpaired populations, not adjusted for multiple comparisons.

Antibody reactivity to the receptor-binding domain (RBD) of the SARS-CoV-2 spike, full-length spike (S), and the nucleocapsid (N) was assayed by IgG/IgA standard enzyme-linked immunosorbent assays (ELISA) and IgG high-throughput MSD V-PLEX assay. An ACE2-SARS-CoV-2 RBD inhibition assay was used to detect neutralising antibodies^5,8^.

All treatments were associated with significantly reduced IgG antibody responses compared to healthy controls for all SARS-CoV-2 antigens, using an MSD V-PLEX assay (Figure 1). The greatest reduction in IgG response by ELISA was observed in individuals treated with infliximab/thiopurine combination therapy (Figure 2a; p=0.00019). Furthermore, IgA responses were significantly reduced in individuals treated with infliximab/thiopurine combination therapy compared to healthy controls (Figure 2b; p=0.009), but not in IBD patients treated with infliximab or vedolizumab monotherapy.

Next, we utilized an ELISA-based inhibition assay to determine the ability of serum to neutralize the binding of SARS-CoV-2 RBD-ACE2 interaction (Figure 2c). Individuals treated with vedolizumab or infliximab monotherapy did not show a significant difference in neutralising antibody responses compared to healthy individuals (Figure 2c). However, individuals treated with infliximab/thiopurine combination therapy showed a significantly reduced response compared to either monotherapy groups, and to the healthy control group (Figure 2c, p=0.0054, 0.0022 and p= 0.0092).

Our data are novel, firstly in demonstrating that infliximab/thiopurine combination therapy is associated with significantly lower IgA as well as a range of IgG responses, and most importantly, with impaired functional neutralising antibody responses, compared to responses in healthy individuals. Secondly, we show that whilst IgG responses were significantly reduced in individuals with IBD treated with infliximab or vedolizumab monotherapy compared to healthy controls, this was not the case for IgA and neutralising antibody responses. As neutralising antibody responses are demonstrated to correlate directly with protection (and inversely with severity^9,10^), this observation may provide the mechanistic explanation for the observation reported by the SECURE-IBD study that individuals with combination therapy were at greater risk of severe COVID-19 outcomes than patients on monotherapy^9,10^.

In demonstrating that these therapeutic interventions are selectively associated with a pattern of attenuated antibody responses to SARS-Cov2 infection compared to healthy controls, we believe these data extend current understanding in this important area, and have potentially important implications for patient care and vaccination strategies.

## Data Availability

All data produced in the present study are available upon reasonable request to the authors

## Supplementary Material

**Supplementary Table 1.**
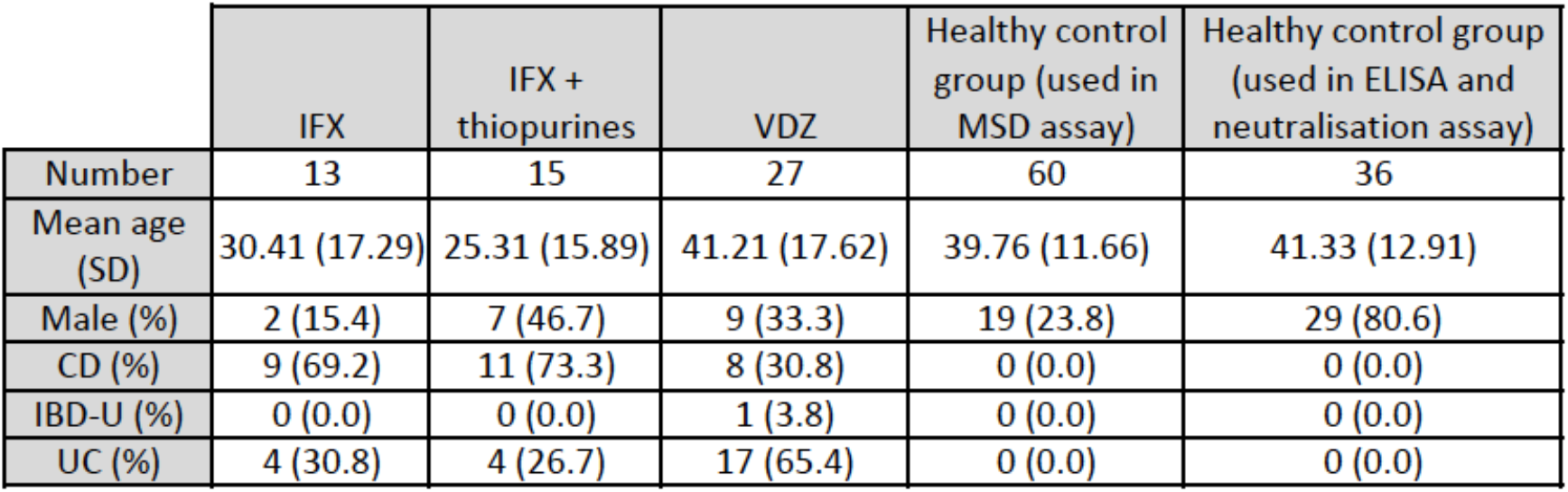
Details of the groups used in the study. Ifx = infliximab, ifx + thiopurines = infliximab plus thiopurines and vdz = vedolizumab. SD = standard deviation. Brackets contain the percentage of the overall total attributed to each group. CD = Crohn’s disease, IBD-U = IBD unclassified, UC = ulcerative colitis.

## Materials and methods

### Setting and participants

Our cohort of 640 IBD patients as previously described by McGregor et al. was followed- up over time until vaccination for COVID-19^1^. This cohort includes adult and paediatric patients from the Royal London Hospital (London, UK) and adult patients from the John Radcliffe Hospital (Oxford, UK). All are IBD patients managed with intravenous anti-TNF therapy (infliximab or biosimilars) or anti-integrin therapy (vedolizumab). Excess serum from routine blood tests was collected at every patient visit. Demographic, socioeconomic, and clinical data were collected through questionnaires and use of the electronic medical record. In order to compare this IBD cohort to a healthy control population, serum samples and data from SARS-CoV-2 PCR positive health care workers of the John Radcliffe Hospital were included in our experimental work and analysis^2^.

### Ethical approval

Samples from Oxford IBD patients were collected as a project (ref: ORB 20/A054) under the ethical approval of the Oxford Radcliffe Biobank, a research tissue bank that has a favourable opinion from the Oxford C South Central REC (ref:19/ SC/0173).

Samples from London IBD patients were collected as a project under the ethical approval of the Digestive Disease Bioresource, Barts Health NHS Trust, a research tissue bank that has a favourable opinion from the Bromley REC, (ref: 15/LO/2127).

Asymptomatic staff sample and data collection (N=36) were part of enhanced hospital infection prevention and control measures instituted by the UK Department of Health and Social Care. Deidentified data from staff testing and patients were obtained from the Infections in Oxfordshire Research Database (IORD) which has generic Research Ethics Committee, Health Research Authority and Confidentiality Advisory Group approvals (19/SC/0403, ECC5-017(A)/2009). Healthy control participants were recruited under the GI Biobank Study 16/YH/0247, approved by the research ethics committee (REC) at Yorkshire & The Humber - Sheffield Research Ethics Committee on 29 July 2016, which has been amended for this purpose on 8 June 2020.

### Enzyme-linked immunosorbent assay (ELISA)

IgG and IgA antibody responses to the spike of SARS-CoV-2 were measured using ELISAs. Nunc-Immuno 96-well plates (Thermo Fisher Scientific, USA) were coated with 1.0 μg ml−1 of antigen in PBS buffer and left overnight at 4 °C. The SARS-CoV-2 spike protein antigen was bought from Native Antigen (UK). Plates were washed with 3x with 0.1% PBS–Tween (PBS/T), then blocked with casein in PBS for 1 hour at room temperature (RT). Serum or plasma was diluted in casein–PBS solution at dilutions ranging from 1:50 to 1:1,500 before being added to NuncImmuno 96-well plates in triplicate. Plates were incubated for 2 hours before being washed with 6x with PBS/T. Secondary antibody rabbit anti-human whole IgG conjugated to alkaline phosphatase (Sigma, USA) or a secondary antibody mouse anti-human IgA conjugated to horse radish peroxidase (Sigma, USA) was added at a dilution of 1:1000 in casein–PBS solution and incubated for 1 hour at RT after which a final wash was performed. For detection of the IgG secondary antibody, plates were developed by adding 4-nitrophenyl phosphate substrate in diethanolamine buffer (Pierce, Loughborough, UK), and optical density (OD) was read at 405 nm using a GloMax (Promega, USA). For detection of the IgA secondary antibody, plates were developed by adding Tetramethylbenzidine (TMB) (Thermo scientific USA) to visualize and Sulfuric acid to stop the reaction. The absorbance was read at 450 nm using GloMax (Promega, USA). A reference standard comprised of pooled highly cross-reactive serum was used on each plate to produce a standard curve.

### MSD V-PLEX assay

IgG antibody responses to SARS-CoV-2 spike, RBD, NTD and nucleocapsid and the spike proteins of SARS-CoV-1, HCoV-229E, HCoV-NL63, HCoV-HKU1 and HCoV-OC43 were assessed using the Meso Scale Diagnostics (MSD) Multi-Spot Assay System (MSD, USA). Pre-coated plates (‘Coronavirus panel 2’) were incubated at RT with Blocker A solution for at least 30 minutes whilst being shaken at 500-700 rpm. Serum or plasma was diluted in Diluent 100 at dilutions of 1:500 to 1:50,000 and samples were added to the plates in duplicate. Plates were incubated for 2 hours at RT, whilst being shaken at 500-700 rpm throughout. A 1x working concentration of the SULFO-TAG anti-human IgG Detection Antibody was prepared in Diluent 100. After incubation with the samples, the plates were washed x3 with 1x MSD Wash buffer. Prepared detection antibody solution was added to the plates, which were incubated at RT for 1 hour, whilst being shaken. Plates were then washed x3 with 1X MSD Wash buffer. To read the assay results, MSD GOLD Read Buffer B (provided ready to use) was added to the plate. No incubation is required, and the plates were read on a MESO QuickPlex SQ 120 (MSD, USA) immediately after adding the buffer. A 7-point calibration curve of the standards was prepared using Diluent 100. Diluent 100 was used as a negative control. An additional three positive controls provided with the kit were also run on every plate. All standards and controls were run in duplicate. Data from the assay was analysed using MSD Discovery Workbench software, which averaged all the duplicates, generated, and fitted the data to standard curves^3^.

### SARS-CoV-2 RBD-ACE2 Inhibition Assay

The qualitative immunoenzymatic determination of RBD-ACE2 inhibition antibodies is based on an ELISA based assay technique (The Native Antigen Company, Oxford, UK). Microplates were coated with RBD in Dulbecco’s Phosphate Buffered Saline (DPBS) to bind corresponding ACE2 or blocking antibodies of the sample. After washing the wells with DPBS + 0.05% Tween 20 to remove all unbound sample material, serum or plasma samples were added at a 1:20 dilution and allowed to bind. After incubation a horseradish peroxidase (HRP) labelled ACE2 conjugate is added and incubated. This conjugate binds to the captured RBD which has not been bound by the antibody sample. In a second washing step, unbound conjugate is removed. Bound ACE2-HRP conjugate (that therefore represents the absence of neutralizing antibody), is visualized by adding TMB substrate. Sulphuric acid was added to stop the reaction. Absorbance at 450nm was then read using a GloMax microplate reader (Promega, USA).

### Statistical analysis

P-values are derived from a Wilcoxon (rank-sum) test for unpaired populations, not adjusted for multiple comparisons.

## Funding

ME was supported by the Helmsley Trust as part of the ICARUS-IBD study [Grant Number 2107–04731]. This healthcare worker study was funded by the UK Department of Health and Social Care as part of the PITCH (Protective Immunity from T cells to Covid-19 in Health workers) Consortium, with contributions from UKRI/NIHR through the UK Coronavirus Immunology Consortium (UK-CIC), the Huo Family Foundation and The National Institute for Health Research (UKRIDHSC COVID-19 Rapid Response Rolling Call, Grant Reference Number COV19-RECPLAS). None of our funding bodies had any role in study design, data acquisition, data analysis or reporting of the results.

## Competing Interests Statement

JS has received lecture fees from Takeda and from the Falk Foundation.

## Notes

### Author Declarations

Samples from Oxford IBD patients were collected as a project (ref: ORB 20/A054) under the ethical approval of the Oxford Radcliffe Biobank, a research tissue bank that has a favourable opinion from the Oxford C South Central REC (ref:19/ SC/0173). Samples from London IBD patients were collected as a project under the ethical approval of the Digestive Disease Bioresource, Barts Health NHS Trust, a research tissue bank that has a favourable opinion from the Bromley REC, (ref: 15/LO/2127). Deidentified data from staff testing and patients were obtained from the Infections in Oxfordshire Research Database (IORD) which has generic Research Ethics Committee, Health Research Authority and Confidentiality Advisory Group approvals (19/SC/0403, ECC5-017(A)/2009). Healthy control participants were recruited under the GI Biobank Study 16/YH/0247, approved by the research ethics committee (REC) at Yorkshire & The Humber - Sheffield Research Ethics Committee on 29 July 2016, which has been amended for this purpose on 8 June 2020.

